# Assessing the impact of variation in diagnostic coding among the three countries in the UK Biobank

**DOI:** 10.1101/2024.12.13.24319003

**Authors:** Lei Clifton, Wenyu Liu, Jennifer A Collister, Thomas J Littlejohns, Raphael R Goldacre, Naomi Allen, David J Hunter

## Abstract

1.

**Background:** The UK Biobank (UKB) study has linked hospital inpatient data collected from England, Scotland, and Wales, which use different clinical coding systems to record health outcomes. Scotland records up to 6 different diagnostic codes for one inpatient episode, compared with up to 20 in England, and 14 in Wales. We assessed the impact of the variations in diagnostic coding among countries on observed disease incidence rates.

**Methods:** We examined the number of diagnoses coded by each country, and then compared the incidence of three diseases between countries: Parkinson’s disease (PD), type 2 diabetes (T2D), and dementia. We constructed Cox models for each disease, adjusting for “country”.

**Results:** England appears to have the highest risk (hazard ratio, HR) for all three diseases, while Scotland appears to have the lowest risk: HR [95% CI] = 0.62 [0.54, 0.72] for PD, 0.49 [0.45, 0.54] for T2D, and 0.88 [0.78, 0.99] for dementia.

**Conclusions:** The observed incidence of these diseases and the estimated effect of “country” in Cox models are likely influenced by the clinical coding variations among countries. Researchers need to be aware of this and account for these variations in their analyses.

**Key messages in a box:** What is already known on this topic:

- There are variations in the clinical coding systems in hospitals among the three countries in UKB.
- These variations could impact research investigating geographic differences in disease incidence. What this study adds:
- We assessed the impact of the variations in diagnostic coding among the three countries in UKB.
- We showed that the observed disease incidence and the estimated effect of “country” are likely influenced by these variations.

How this study might affect research, practice or policy:

- Researchers need to understand the provenance of data and account for the different diagnostic coding and other geographic variation specific to each of three countries, in order to reach robust conclusions that are not influenced by artefacts.

## 3. Introduction

Routinely collected health statistics in the United Kingdom play an important role in population health research, health resource allocation, and descriptive and observational epidemiology. A major data source is hospital inpatient data that contains information about reasons for hospitalization and additional diagnoses not necessarily related to the hospitalization. These hospital inpatient data are recorded in the Hospital Episode Statistics (HES) for England, Scottish Morbidity Record (SMR), and Patient Episode Database for Wales (PEDW). We will collectively refer to all these three sources of hospital inpatient data as Admitted Patient Care (APC). The disease diagnostic (e.g. ICD-10) codes in APC are currently the main source of outcome ascertainment for researchers. In all three countries, admitted episodes of patient care are assigned one main (primary) code for the reason for admission, as well as a variable number of secondary (or underlying) disease codes. The primary diagnosis in APC is recorded at “location 0”, with secondary diagnoses recorded at “location 1, 2, 3 …”. However, Scotland records a maximum of 6 primary plus secondary diagnostic codes for one episode, compared with up to 20 in England, and 14 in Wales. In England the number of diagnosis fields has increased over time, from up 7 codes before April 2002, up to 14 in April 2002 - March 2007, and up to 20 since April 2007 [1].

This variability in the coding practices for the APC data by country may lead to variation in the likelihood of conditions being recorded that are not the main (primary) reason for an admitted episode and yield mistaken inferences about the relative burden of diseases across countries. We sought to understand the likely impact on three diseases that are more likely to be recorded as a secondary diagnosis: Parkinson’s disease (PD), type 2 diabetes (T2D), and all-cause dementia.

The UK Biobank (UKB) is an ongoing prospective cohort study of approximately 500,000 participants recruited from 22 assessment centres across England, Scotland, and Wales between 2006 and 2010 [2]. As primary care records are currently only available for approximately half of the cohort participants for a limited period of follow-up, the APC records are used in most analyses to identify incident disease. The linkage to electronic hospital records in UKB offered the opportunity to evaluate whether differences in coding practices in the APC data between the three countries influence conclusions about regional variation in disease incidence. To the best of our knowledge, there have been no studies examining the impact of this variability.

## 4. Methods

We first examined the distribution of the number of “primary + secondary” ICD-10 diagnostic records per episode in the APC records from the three countries, excluding duplicates within each episode and truncated at the earliest hospital inpatient data censoring date across all data providers (31 May 2022).

For each individual, we defined “the first disease location” as the first occurred location of the disease diagnostic codes among all episodes of a patient. Within each episode, we first find the first occurred location among all the disease diagnoses, since a specific disease may be defined by several ICD codes. Since multiple ICD codes can represent a single disease, the first recorded ICD code for that disease was used. We then take the minimum location of these first locations among all episodes, since a patient may have multiple hospital stays which may contain multiple episodes. This method yielded, for each patient, a number for the “first disease location” for each disease.

For example, if dementia is the primary diagnosis (i.e. the main reason for admission) in any episode of an individual, their “first disease location” for dementia will be “location 0”. If a disease dementia is first recorded as a secondary diagnosis in an episode and then became a primary diagnosis (at location 0) in a later episode, the “first disease location” for dementia of that person will still be “location 0”. If the disease was never recorded as the primary diagnosis, the “first disease location” can be at “location 1, 2, 3 …”, whichever is the first occurred location across all episodes for that individual.

For PD and dementia, we used the outcome definition derived by the UKB (<u>Resource 594</u>). T2D related diagnostic code list is currently not available from UKB. We instead used clinical codes as reported from the existing literature [3]. For description of analysis populations and outcome definitions, please see Supplementary Section 3.

We then constructed Cox proportional hazards models to quantify and compare the effect of “country” alongside established risk factors on the risk prediction of each disease. We excluded prevalent cases at baseline, using APC and UKB self-report data. For incident cases in our disease prediction Cox model, we used both APC and Death Registry for accurate disease ascertainment and risk. We included established disease-specific risk factors available in UKB, and an additional covariate “country”, in the Cox model for each disease [4].

## 5. Results

Of 502,358 participants, 444,908 have at least one hospital inpatient record, with 11%, 15%, and 12% participants, in England (N=445,718), Scotland (N=35,836) and Wales (N=20,804), respectively, not appearing in the APC data over the follow-up period. Baseline characteristics by country are provided in Table S1. The prevalent and incident cases for each disease are shown in Tables S3 and S5 – S7, with incident rates shown in Table S4.

Across all diseases recorded in APC, Scotland has a notably higher percentage (37%) of episodes with one diagnostic code (i.e. primary diagnosis only) than England and Wales (both approximately 23%, Figure 1 top). For episodes with six diagnostic codes (five of which are secondary diagnoses), Scotland again recorded a much higher percentage (11%) than those in England and Wales (both at approximately 7%). In England and Wales, 19.2% and 17.8% of diagnoses are recorded after the first 6 diagnostic codes, respectively. The median number of ICD diagnostic codes per episode is 3 for England and Wales, compared with 2 for Scotland.

**Figure 1.**
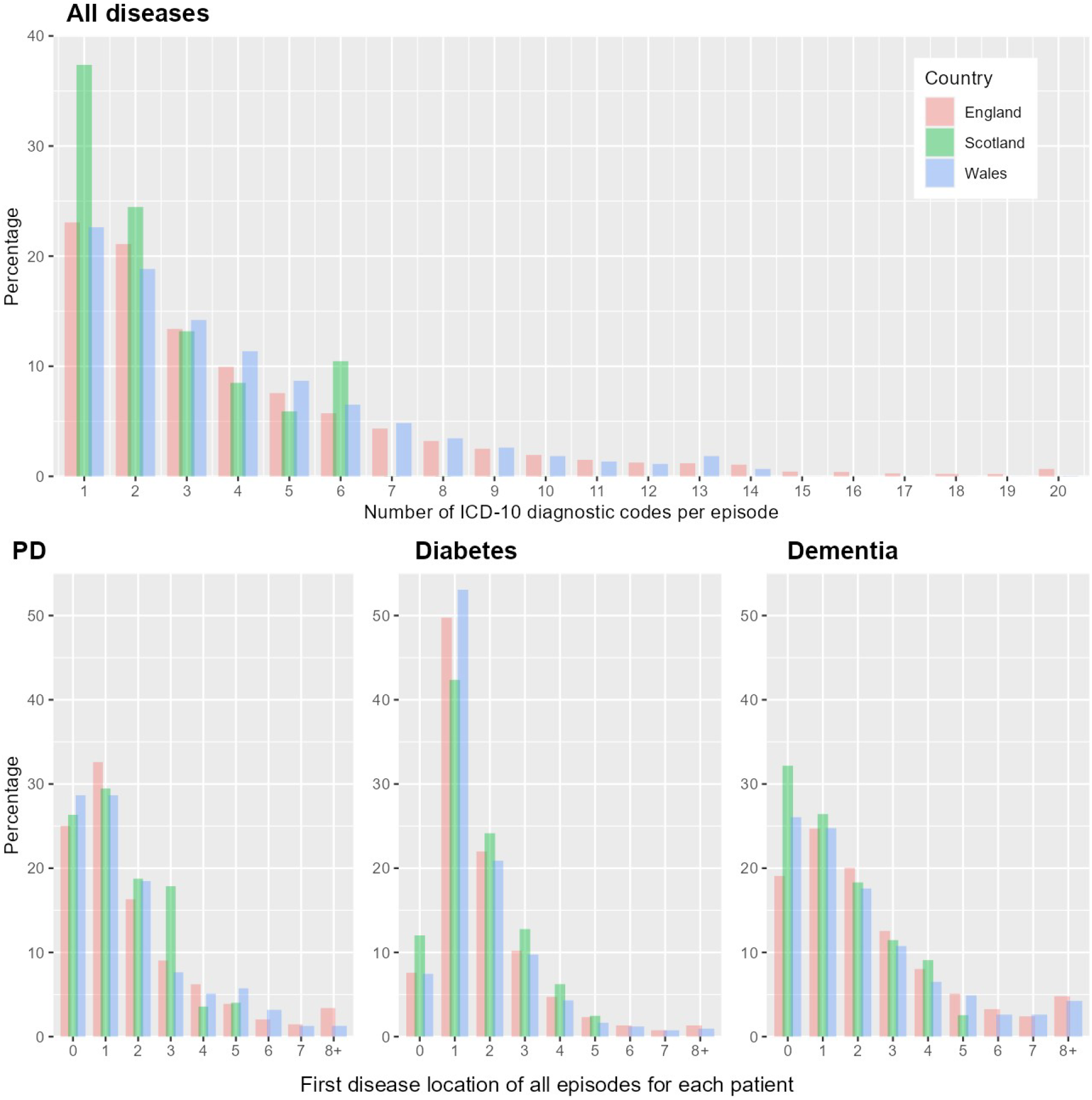
Top: Number of “primary + secondary” ICD-10 diagnostic codes per episode, excluding duplicates within each episode and truncated at the earliest hospital inpatient data censoring date among countries. England records up to 20 diagnostic codes per episode, Scotland up to 6, and Wales up to 14. The sum of all counts is the number of episodes (not patients). For the raw number of ICD-10 diagnostic codes per episode by country, please see Table S2. The median number of ICD-10 diagnostic codes per episode is 3 for England and Wales, compared with 2 for Scotland. Detailed figures by age and sex are shown in Figures S1a-S1c. Bottom: First disease location of all available episodes for each patient with Parkinson’s disease, diabetes and dementia, respectively, by country. Scotland appears to have different recording patterns from England and Wales for PD and diabetes, but less so for dementia. We note that the disease location ranges from 0 to 19 for England (i.e., max 20 diagnostic codes per episode), from 0 to 5 for Scotland (i.e. max 6 diagnostic codes) and up to 13 for Wales (i.e. max 14 diagnostic codes), where primary diagnoses are recorded as “location 0” in the raw data. For brevity, the bottom figures show “8+” disease location.

In England, Scotland and Wales, respectively, the percentage of primary diagnoses is 25.0%, 26.3%, 28.7% for PD; 7.6%, 12.0%, 7.5%, for T2D; and 19.1%, 32.2%, 26.1% for dementia (Figure 1 bottom). T2D is less likely to be recorded as the primary diagnosis in all three countries, especially in England and Wales, compared with PD and dementia. Dementia is more likely to be recorded as the primary diagnosis in Scotland than England and Wales, compared with PD and T2D. The pattern of the “first disease location” for PD is similar in all three countries.

After applying the exclusion criteria (Figures S2 - S4) to the 502,358 UKB participants, approximately 499,000, 473,000, and 204,000 participants are included in the study populations for PD, T2D and dementia, respectively. The dementia population is restricted to individuals aged 60-70 years, as they are at greatest risk of developing dementia. Baseline characteristics are shown in Tables S8 – S10. Table 1 shows the estimated effect (hazard ratios, HR) of variable “country” obtained from Cox model after adjusting for known risk factors for each specific disease. For all three diseases, England (the reference category) has the highest risk, while Scotland has the lowest risk with HR = 0.62, 0.49, 0.88, for PD, T2D, and Dementia, respectively. These lower risks in Scotland are potentially misleading, as they may be influenced by the coding variation across countries, especially for secondary conditions. The variable “country” is statistically significant (p-value < 0.0001) for PD and T2D, but not for dementia. One possible explanation is that compared with PD and T2D, dementia may be regarded as an important condition to be recorded in earlier locations of diagnoses and Scotland had the highest proportion of dementia as a primary diagnosis. The HR of established risk factors for the three disease outcomes was not materially affected when adjusted for “country” (Tables S11 – S13). The proportional hazards assumption was met by visually assessing the scaled Schoenfeld residuals via the “survminer” R package.

**Table 1:** Hazard ratio (HR) of variable “country” obtained from Cox model after adjusting for known risk factors for each disease^1^. HR is shown with its associated 95% confidence interval (CI). PD: Parkinson’s Disease. T2D: Type 2 Diabetes. p-value: overall p-value of variable “country”. Detailed results are in supplementary Tables S11 - S13.

|  | England | Scotland | Wales | p-value |
| --- | --- | --- | --- | --- |
| **PD** | **(N = 442,577)** | **(N = 35,614)** | **(N = 20,739)** |  |
|  | 1 (reference) | 0.62 [0.54, 0.72] | 0.77 [0.64, 0.92] | <0.0001 |
| **T2D** | **(N = 419,839)** | **(N = 34,014)** | **(N = 19,612)** |  |
|  | 1 (reference) | 0.49 [0.45, 0.54] | 0.96 [0.89, 1.04] | <0.0001 |
| **Dementia** | **(N = 181,850)** | **(N = 13,773)** | **(N = 7,983)** |  |
|  | 1 (reference) | 0.88 [0.78, 0.99] | 0.98 [0.83, 1.16] | 0.098 |

<sup>1</sup>Established risk factors included for PD: age, sex, self-reported ethnicity, Townsend deprivation score quintile, family history of PD, and smoking.
T2D: age, self-reported ethnicity, Townsend deprivation score quintile, family history of diabetes, waist circumference (cm), BMI, physical inactivity, and hypertension.
Dementia: age, sex, self-reported ethnicity, Townsend deprivation score quintile, family history of dementia, smoking, APOE\_e4\_carrier, education, physical inactivity, depression, diabetes, hearing impairment, hypertension, social isolation, and BMI.

## 6. Discussion

The main strength of our study is the systematic examination using the large UKB cohort. One limitation is the relatively small numbers of participants in Wales and Scotland, compared with England. Nevertheless, the availability of in-hospital patient data across UKB has enabled us to examine the impact of disease coding variation among the three countries.

In general, Scotland displayed different recording patterns for inpatient diagnostic codes, compared with England and Wales. This is more prominent for PD and T2D, than dementia. Variation in disease coding across jurisdictions in any study that analyses data across these jurisdictions may influence the interpretation of studies seeking to understand geographic differences in disease incidence. Here we demonstrate that differences in the maximum number of ICD codes routinely recorded in hospital inpatient records in England, Scotland, and Wales may be misinterpreted as differences in the incidence rates of a disease between these countries. For instance, a recent publication [5] using UKB data reported lower risk of heart failure in urban Scotland than England. The authors speculated that Scottish population is densely concentrated in Glasgow and Edinburgh in the central lowlands, thus potentially benefiting from easier access to health services. Our study provides another plausible explanation (i.e. coding variation across countries) for this phenomenon.

Similarly, a recent study [6] examining health disparities in the United Kingdom using the UKB database reported a higher relative burden of disease in England than Scotland and Wales. The authors postulated that this may be attributed to higher social economic deprivation (i.e. lower social economic status) for participants recruited from England compared to those from Scotland and Wales. However, we observed that 23.9% of UK Biobank participants in Scotland were in the most deprived quintile (measured by the Townsend Deprivation index) compared with 19.9% of participants in England). A plausible explanation for their findings is the systematically lower incidence cases for PD and T2D (but not for dementia) in Scotland due to the smaller number of conditions coded. Studies using data that is geographically determined such as air pollution exposure, or latitude, could be biased if geographical differences in disease ascertainment are not accounted for.

We examined this in the UK Biobank, but the same issue would apply to any study examining incidence rates across these three countries. Although this issue is clearly flagged in the UK Biobank documentation, it could be easily overlooked.

In the present analysis, we cannot exclude the possibility that differences in the health systems in the devolved nations in terms of how patients are managed at home or in a care home, might also influence rates of hospitalisation and thus recording of diagnoses. However, the differences in the number of diseases coded plausibly explain a large proportion of the differences in incident cases.

The increasing availability of large databases that can be used for both epidemiologic and health care policy studies has led to an explosion in publications in both domains. In addition to geographical variation in disease coding we have previously noted [4] that for conditions that are not often the principal cause of a hospitalization that there is often a lag of many years before a diagnosis recorded in the primary care record appears in the APC data, leading to an underestimation of incidences and age at onset of these conditions. Thus, those analysing these databases need to take the time to understand the provenance of the data and rules that govern these databases in order to reach robust conclusions that are not influenced by artefacts and idiosyncrasies in the recording of the data.

## 7. Conclusion

Scotland appears to have the lowest risk (measured by HR) of PD, T2D, and dementia, among the three countries. This is likely misleading, probably influenced by the coding variation (owing to fewer opportunities to record secondary diagnoses in hospital data from Scotland) and potentially other unobserved differences in clinical management between the three countries.

## Data Availability

All data produced in the present work are contained in the manuscript

https://www.ukbiobank.ac.uk

## 8. Declarations

### 8.1 Ethics approval and consent to participate

The UK Biobank study received ethical approval from the North West Multi-centre Research Ethics Committee (REC reference: 11/NW/03820). All participants gave written informed consent before enrolment in the study, which was conducted in accordance with the principles of the Declaration of Helsinki. Patient and community involvement

The analyses presented here are based on existing data from the UK Biobank cohort study, and the authors were not involved in participant recruitment. No patients were asked to advise on interpretation or writing these results. Results from UK Biobank are routinely disseminated to study participants via the study website and social media outlets.

### 8.2 Consent for publication

Yes.

### 8.3 Availability of data and material

This research has been conducted using the UK Biobank Resource under Application Number 33952. Requests to access the data should be made via application directly to the UK Biobank, *https://www.ukbiobank.ac.uk*

The code used for analyses are available at https://gitlab.com/wenyul/countrydiff

### 8.6 Authors’ contributions

LC and DJH conceived the project; LC and WL outlined the statistical methods; LC, WL, and DJH drafted the manuscripts; WL conducted the statistical analysis; JAC reviewed the R scripts by WL. All authors have contributed to the study design, revised the manuscript, and agreed on its contents.

### 8.7 Transparency statement

The lead author affirms that this manuscript is an honest, accurate, and transparent account of the study being reported; that no important aspects of the study have been omitted; and that any discrepancies from the study as planned have been explained.

## 8.8 Acknowledgements

We thank the participants and staff of the UK Biobank for enabling us to conduct this research. This study has been conducted using the UK Biobank Resource under Application Number 33952.

