## Supplements for "Assessing the impact of variation in diagnostic coding among the three countries in the UK Biobank"

### 1. Summary statistics including all diseases

#### 1.1 Baseline characteristics

The baseline assessment of UKB was performed in-person, while follow-up outcomes are obtained via linkage to electronic records, including hospital inpatient records, cancer and death registry data.

Table S1: Baseline characteristics of all UKB participants in England, Scotland, and Wales. It includes the whole UKB population (showing column percentage %), before applying any exclusion criteria for specific diseases.

|  | **England (N=445,718)** | **Scotland (N=35,836)** | **Wales (N=20,804)** | **Total (N=502,358)** |
| --- | --- | --- | --- | --- |
| **Baseline Age (years)** |  |  |  |  |
| Mean (SD) | 57.06 (8.10) | 56.83 (8.07) | 56.66 (7.92) | 57.03 (8.09) |
| Min, Max | 37.42, 73.69 | 40.34, 70.90 | 40.25, 70.96 | 37.42, 73.69 |
| **Ethnicity** |  |  |  |  |
| White | 417,262 (93.6%) | 35,124 (98.0%) | 20,178 (97.0%) | 472,564 (94.1%) |
| Black | 7,906 (1.8%) | 57 (0.2%) | 95 (0.5%) | 8,058 (1.6%) |
| S. Asian | 7,720 (1.7%) | 185 (0.5%) | 116 (0.6%) | 8,021 (1.6%) |
| Mixed | 2,727 (0.6%) | 90 (0.3%) | 136 (0.7%) | 2,953 (0.6%) |
| Other | 7,521 (1.7%) | 267 (0.7%) | 197 (1.0%) | 7,985 (1.6%) |
| Missing | 2,582 (0.6%) | 113 (0.3%) | 82 (0.4%) | 2,777 (0.6%) |
| **Sex** |  |  |  |  |
| Female | 242,065 (54.3%) | 19,942 (55.6%) | 11,287 (54.3%) | 273,294 (54.4%) |
| Male | 203,653 (45.7%) | 15,894 (44.4%) | 9,517 (45.7%) | 229,064 (45.6%) |
| **Townsend Deprivation Index Quintile** |  |  |  |  |
| Q1: least deprived | 88,557 (19.9%) | 8,043 (22.4%) | 4,717 (22.7%) | 101,317 (20.2%) |
| Q2 | 90,310 (20.3%) | 6,006 (16.8%) | 4,273 (20.5%) | 100,589 (20.0%) |
| Q3 | 88,159 (19.8%) | 6,232 (17.4%) | 4,770 (22.9%) | 99,161 (19.7%) |
| Q4 | 89,402 (20.1%) | 6,937 (19.4%) | 4,145 (19.9%) | 100,484 (20.0%) |
| Q5: most deprived | 88,770 (19.9%) | 8,543 (23.8%) | 2,869 (13.8%) | 100,182 (19.9%) |
| Missing | 520 (0.1%) | 75 (0.2%) | 30 (0.1%) | 625 (0.1%) |
| **Family History of PD** |  |  |  |  |
| No family history | 427,968 (96.0%) | 34,520 (96.3%) | 19,947 (95.9%) | 482,435 (96.0%) |
| Family history | 17,750 (4.0%) | 1,316 (3.7%) | 857 (4.1%) | 19,923 (4.0%) |
| **Family History of Diabetes** |  |  |  |  |
| No family history | 348,561 (78.2%) | 29,165 (81.4%) | 16,081 (77.3%) | 393,807 (78.4%) |
| Family history | 97,157 (21.8%) | 6,671 (18.6%) | 4,723 (22.7%) | 108,551 (21.6%) |
| **Family History of Dementia** |  |  |  |  |
| No family history | 393,532 (88.3%) | 31,709 (88.5%) | 18,703 (89.9%) | 443,944 (88.4%) |
| Family history | 52,186 (11.7%) | 4,127 (11.5%) | 2,101 (10.1%) | 58,414 (11.6%) |

#### 1.2 Number of “primary + secondary” ICD-10 diagnostic codes per episode

Table S2 below shows the number of ICD-10 diagnostic codes per episode by country, corresponding to Figure 1 (Top) in the main paper. Note that a patient stay may contain multiple episodes.

*Table S2. Number of distinct primary + secondary diagnostic codes per episode by country, censored at the earliest UKB administrative censoring date among the three countries (31 May 2022). We removed duplicate diagnostic codes within an episode. This table corresponds to histogram “number of ICD-10 diagnostic codes per episode” (Figure 1) shown in the main paper. England records up to 20 diagnostic codes per episode, Scotland up to 6, and Wales up to 14.*

| **Number of diagnostic codes per episode** | **England** | | **Scotland** | | **Wales** | |
| --- | --- | --- | --- | --- | --- | --- |
|  | **Count** | **Percentage** | **Count** | **Percentage** | **Count** | **Percentage** |
| **1** | 835790 | 23.06 | 102408 | 37.37 | 40087 | 22.62 |
| **2** | 764539 | 21.1 | 67017 | 24.46 | 33381 | 18.83 |
| **3** | 485416 | 13.39 | 36108 | 13.18 | 25170 | 14.2 |
| **4** | 360745 | 9.95 | 23270 | 8.49 | 20134 | 11.36 |
| **5** | 273859 | 7.56 | 16160 | 5.9 | 15377 | 8.68 |
| **6** | 207570 | 5.73 | 28669 | 10.46 | 11537 | 6.51 |
| **7** | 157242 | 4.34 |  |  | 8582 | 4.84 |
| **8** | 116423 | 3.21 |  |  | 6112 | 3.45 |
| **9** | 91109 | 2.51 |  |  | 4627 | 2.61 |
| **10** | 70138 | 1.94 |  |  | 3259 | 1.84 |
| **11** | 54193 | 1.5 |  |  | 2389 | 1.35 |
| **12** | 45158 | 1.25 |  |  | 2001 | 1.13 |
| **13** | 43304 | 1.19 |  |  | 3256 | 1.84 |
| **14** | 38535 | 1.06 |  |  | 1182 | 0.67 |
| **15** | 15752 | 0.43 |  |  |  |  |
| **16** | 14450 | 0.4 |  |  |  |  |
| **17** | 9918 | 0.27 |  |  |  |  |
| **18** | 8008 | 0.22 |  |  |  |  |
| **19** | 7489 | 0.21 |  |  |  |  |
| **20** | 24338 | 0.67 |  |  |  |  |

#### 1.3 Number of “primary + secondary” ICD-10 diagnostic codes per episode by age and sex

Further to Figure 1 (Top) in the main paper, Figures S1a-S1c show the number of ICD-10 diagnostic codes per episode split by age and sex. We observe that the number of ICD-10 diagnostic codes per episode increased with age, while maintaining similar distributions between male and female.

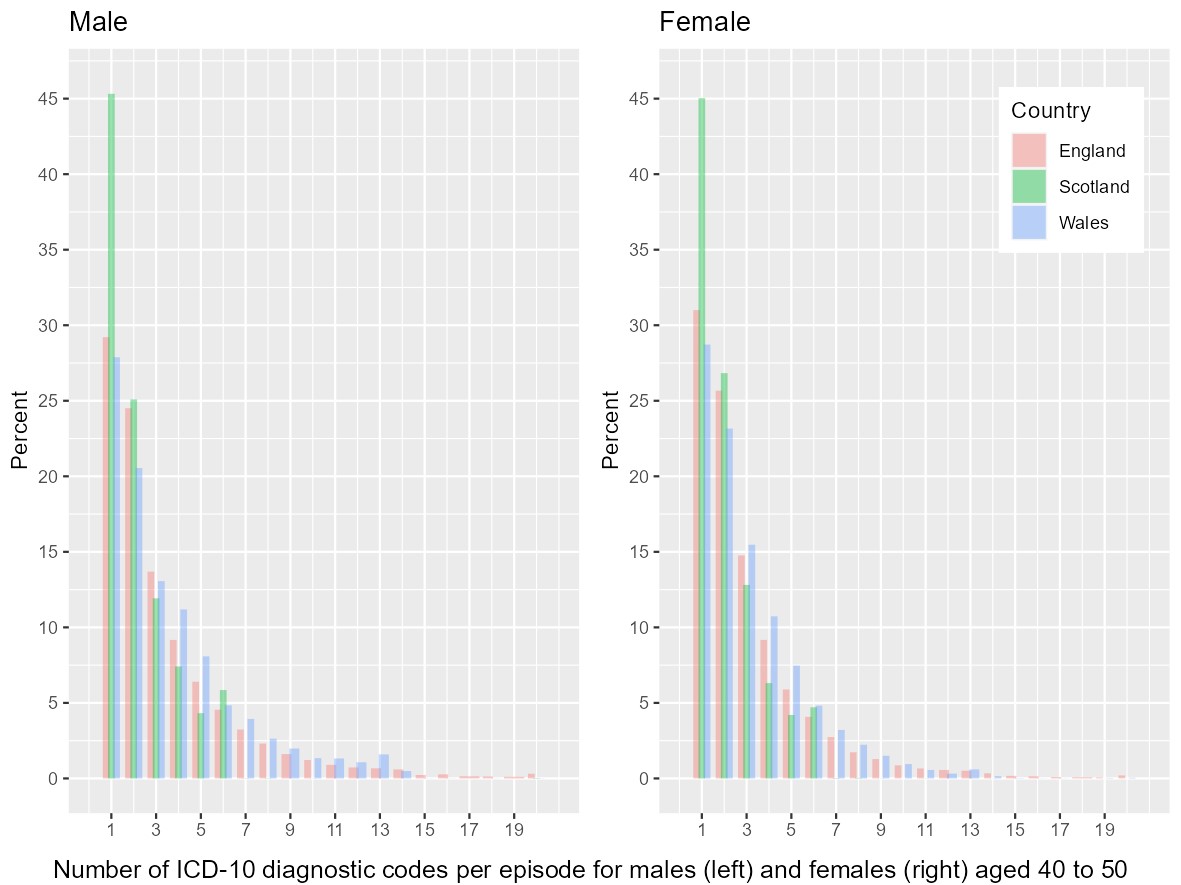

Figure S1a. Number of “primary + secondary” ICD-10 diagnostic codes per episode (excluding duplicates) by sex and country for participants aged [40, 50). The y-axis shows the number/percentage of diagnostic codes (not participants). There are 42,093 men and 55,904 women in age group [40, 50) years.

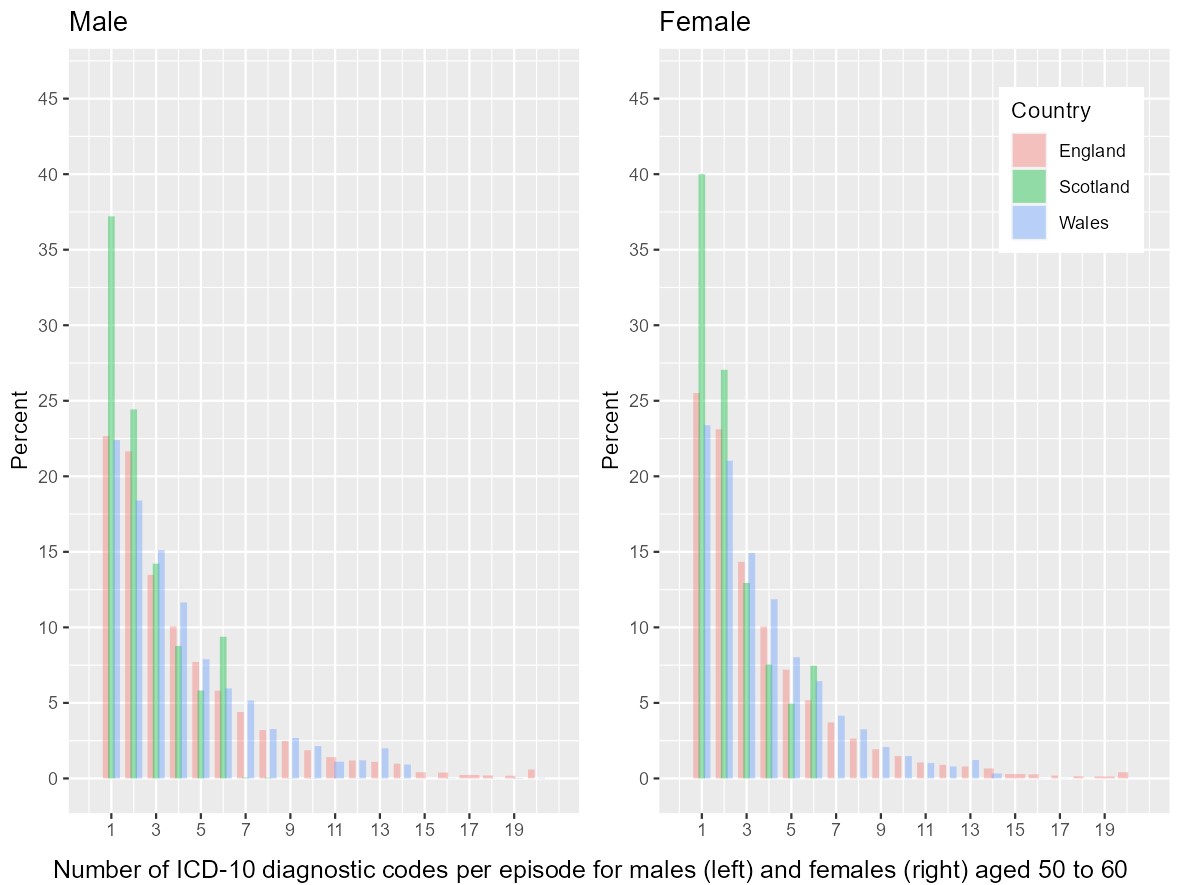

Figure S1b. Number of “primary + secondary” ICD-10 diagnostic codes per episode (excluding duplicates) by sex and country for participants aged [50, 60), including 62,767 men and 81,278 women.

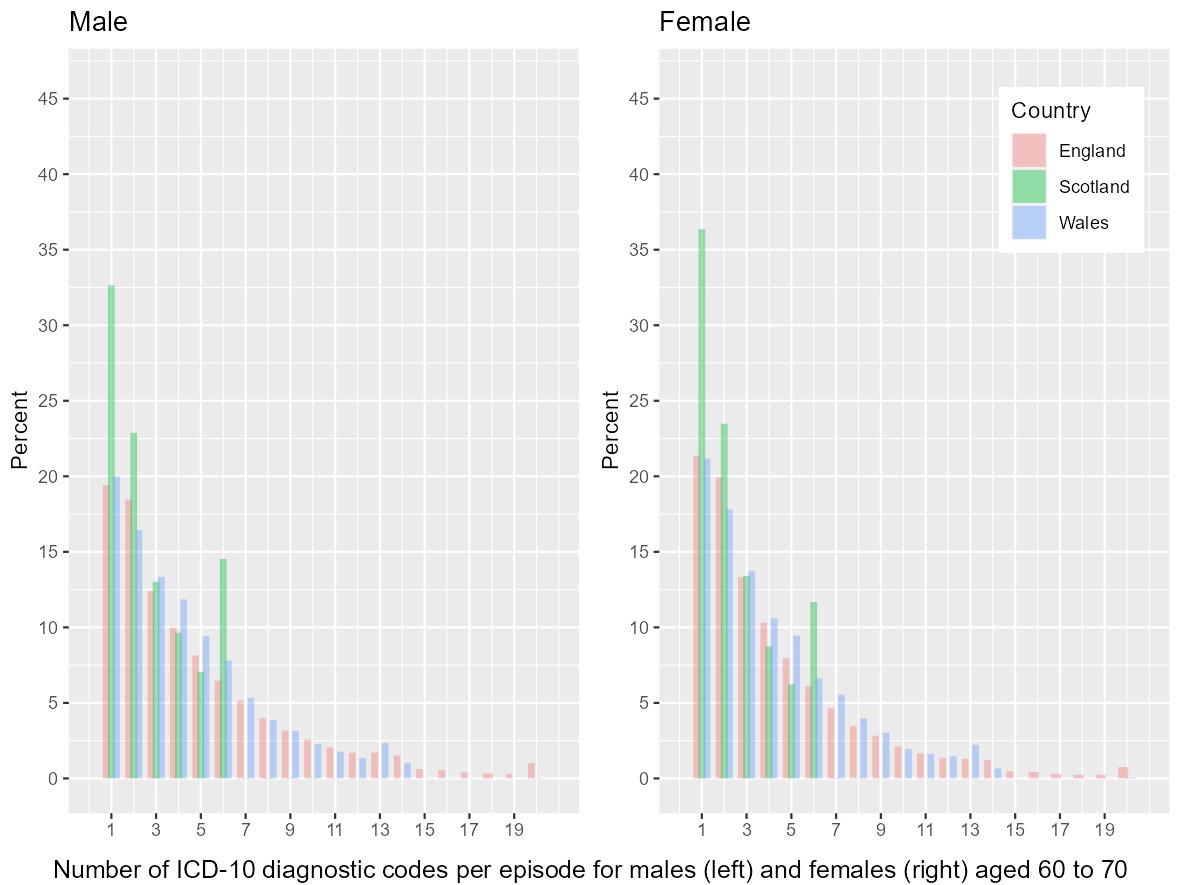

Figure S1c. Number of “primary + secondary” ICD-10 diagnostic codes per episode (excluding duplicates) by sex and country for participants aged [60, 70), including 95,134 men and 105,335 women.

#### 1.4 Number of episodes per spell

Another coding difference across the three countries regards “episodes” and “spell”. According to UKB Resource 138483, in England and Wales, a continuous period in hospital from admission to discharge is called a “spell”, within which there can be multiple “episodes” indicating care under different consultants. In contrast, the episodes of care in Scotland are not arranged into spells in this way. In this paper, we examined diseases by “episode”, instead of “spell” in Admitted Patient Care (APC). One reason for our choice is the guidance from the UKB document (Appendix A, Resource 138483) that states “*the ‘spell_index’ data-field cannot always be relied upon to fully group episodes into spells. Researchers particularly interested in hospital spells should treat these fields with caution, and we recommend you develop your own rules for assigning episodes to spells.*”

The number of episodes per spell (using data-field ‘spell_index’ as a proxy) reveals that most spells consist of only one episode (94.04% in England, 99.95% in Scotland, and 99.78% in Wales); therefore the number of ICD-10 diagnostic codes per “*spell”* is almost identical to that by “episode”.

### 2. Summary statistics of three specific diseases

#### 2.1 Prevalent and incident cases in APC

We identified prevalent and incident cases from multiple sources: UKB self-report and APC for prevalent cases; APC and Death registry for incident cases for accurate disease ascertainment. We note that cases recorded in different sources are not mutually exclusive. The variation in clinical coding between the three countries is specific to APC data, and therefore here we focus on the number and percentage of cases in APC data in Table S3. It shows that the disease percentages in Scotland are lower than England and Wales for PD and T2D, but not so for dementia.

*Table S3: Prevalent and incident cases recorded in APC for each disease across countries. Each cell shows the number of cases (%), where percentage = number of cases / N in each country.*

|  |  | **England (N=445,718)** | **Scotland (N=35,836)** | **Wales (N=20,804)** |
| --- | --- | --- | --- | --- |
| **Prevalent cases** | **PD**^1^ | 367 (0.1%) | 23 (0.1%) | 21 (0.1%) |
|  | **T2D**^2^ | 10,091 (2.3%) | 573 (1.6%) | 499 (2.4%) |
|  | **Dementia**^3,4^ | NA | NA | NA |
| **Incident cases** | **PD**^1^ | 3,953 (0.9%) | 210 (0.6%) | 137 (0.7%) |
|  | **T2D**^2^ | 32,017 (7.2%) | 1,500 (4.2%) | 1,488 (7.2%) |
|  | **Dementia**^3,4^ | 8,138 (1.8%) | 650 (1.8%) | 304 (1.5%) |

^1^Parkinson’s disease (PD) was based on the code lists for “algorithmically-defined outcomes” ([UKB Resource 460](https://biobank.ndph.ox.ac.uk/showcase/refer.cgi?id=460)).

^2^Type 2 Diabetes (T2D) was based on Eastwood (2016) Algorithm 1.

^3^Dementia was based on the code lists for “algorithmically-defined outcomes” ([UKB Resource 460](https://biobank.ndph.ox.ac.uk/showcase/refer.cgi?id=460)), where the self-reported dementia was under the group of Dementia/Alzheimers/Cognitive Impairment.

^4^For dementia, the number of patients aged 60 or above are: N = 194,490 in England; N = 14,658 in Scotland; and N = 8,401 in Wales.

For APC data, UK Biobank Resource 138483 (p.3) states that “*Records date back to 1997 for England, 1998 for Wales and 1981 for Scotland*”. Since Scotland and Wales only account for a small proportion of APC data, we consider all hospital data from the earliest start date for each country to the earliest administrative censoring date (31 May 2022) among the three countries to include as much data as possible. For each of the three diseases, we calculated both the number and percentage of cases for each disease[1]. The number of cases include both prevalent and incident cases, for the purpose of examining all existing disease cases throughout the UKB study period.

#### 2.2 Age- and sex- adjusted incident rate by country

Table S4 below shows crude incident rate, and those adjusted for age only, sex only, and “age + sex”, respectively, by country.

*Table S4: Crude and age- and sex- adjusted incident rates (estimated using Poisson regression) per 100,000 person-year by country for PD, T2D and Dementia*

|  | **Incident rate** | **England** | **Scotland** | **Wales** |
| --- | --- | --- | --- | --- |
| **PD** |  | **(N = 442,577)** | **(N = 35,614)** | **(N = 20,739)** |
|  | **Crude rate** | 60.6 | 39.3 | 43.7 |
|  | **Adjusted for:** |  |  |  |
|  | age only | 61.6 | 41 | 47.3 |
|  | sex only | 60.7 | 39.8 | 43.8 |
|  | age + sex | 61.9 | 41.7 | 47.3 |
| **T2D** |  | **(N = 419,839)** | **(N = 34,014)** | **(N = 19,612)** |
|  | **Crude rate** | 369.6 | 172.1 | 363.8 |
|  | **Adjusted for:** |  |  |  |
|  | age only | 372.2 | 175.2 | 374 |
|  | sex only | 370.5 | 173.7 | 364.8 |
|  | age + sex | 373.2 | 176.9 | 374.9 |
| **Dementia**^1^ |  | **(N = 181,850)** | **(N = 13,773)** | **(N = 7,983)** |
|  | **Crude rate** | 269.1 | 300.7 | 257.1 |
|  | **Adjusted for:** |  |  |  |
|  | age only | 272.2 | 293.2 | 271.6 |
|  | sex only | 269.4 | 302.3 | 257.5 |
|  | age + sex | 272.6 | 294.9 | 271.8 |

^1^The study population for dementia is restricted to 60-70 years of age who are most at risk of developing dementia during the follow-up period.

Note that these age- and sex- adjusted incident rates do not take account of other confounders, and therefore may overestimate or underestimate of the actual associations between exposures and the outcome. These incident rates may even be in the opposite direction of the adjusted hazard ratio (shown in Table 1 of the main paper).

The incident rates in Table S4 and the percentages in Table S3 are not directly comparable. The incident rates in Table S4 are based on the study population for the Cox models (shown in Section 3.1), where the exclusion criteria are applied, and only incident cases are included. In contrast, Table S3 shows the prevalent and incident cases and percentages recorded in impatient data by country for each disease, without applying any exclusion criteria.

#### 2.3 First six diagnostic codes

To take account of the different number of diagnostic codes in APC data among the three countries, we truncated the number diagnostic codes in England and Wales to be six (the same as Scotland), and then compared the disease prevalence before-and-after the truncation. Tables S5 – S7 below show that this slightly reduced the number of cases and the corresponding percentage. We note that the choice of the first six diagnostic codes is influenced by the maximal number allowed in records; therefore, the caveat remains that this simulated comparison does not fully represent the hypothetical situation in which England and Wales record six codes only.

Our proposed “the first disease location” is the minimal location of a disease diagnosis among all episodes. It is not the disease location of the first episode (i.e. the earliest diagnosis). For example, if a disease is first recorded as a secondary diagnosis (i.e. at location 1 onwards) in an episode, and then became a primary diagnosis (at location 0) in a later episode, the first disease location will be 0, indicating a primary diagnosis has been assigned to this disease.

Table S5. Parkinson's disease diagnostic cases and percentage by country, counting each disease once using the first disease location. In Column “First 6 codes only”, we only kept the first six diagnostic codes for England and Wales, for further comparison with Scotland where only up to six diagnostic codes are allowed within one episode. Disease cases include both prevalent and incident cases in APC.

| **Parkinson’s disease** | **All diagnostic codes** | | **First 6 codes only** | |
| --- | --- | --- | --- | --- |
|  | **Cases** | **Percentage** | **Cases** | **Percentage** |
| **England**  (N= 445,718) | 4114 | 0.92% | 3830 | 0.86% |
| **Scotland**  (N= 35,836) | 224 | 0.63% | 224 | 0.63% |
| **Wales**  (N= 20,804) | 157 | 0.75% | 148 | 0.71% |

Table S6. Type 2 diabetes diagnostic cases and percentage by country, counting each disease once using first disease location. Disease cases include both prevalent and incident cases in APC

| **Diabetes** | **All diagnostic codes** | | **First 6 codes only** | |
| --- | --- | --- | --- | --- |
|  | **Cases** | **Percentage** | **Cases** | **Percentage** |
| **England**  (N= 445,718) | 41,705 | 9.36% | 40,276 | 9.04% |
| **Scotland**  (N= 35,836) | 2,021 | 5.64% | 2,021 | 5.64% |
| **Wales**  (N= 20,804) | 2,001 | 9.62% | 1,943 | 9.34% |

Table S7. Dementia diagnostic cases and percentage by country, counting each disease once using first disease location. Disease cases include both prevalent and incident cases in APC

| **Dementia** | **All diagnostic codes** | | **First 6 codes only** | |
| --- | --- | --- | --- | --- |
|  | **Cases** | **Percentage** | **Cases** | **Percentage** |
| **England**  (N= 445,718) | 7,631 | 1.71% | 6,831 | 1.53% |
| **Scotland**  (N= 35,836) | 628 | 1.75% | 628 | 1.75% |
| **Wales**  (N= 20,804) | 307 | 1.48% | 278 | 1.34% |

### 3. Risk prediction for specific diseases

We excluded prevalent case at baseline, and used incident cases for risk prediction. Missing data were imputed by multiple imputation (10 imputed datasets) under the assumption of missing at random using the R software *mice* package version 3.16.0. The missing percentage of all variables are reported in Tables S1, S8-S10. Exclusion criteria of the study population for each of the three diseases are in Figures S2-S4.

#### 3.1 Study populations

The flowcharts below illustrate the study population for the Cox model of each of the three diseases. For dementia, we excluded participants younger than 60 years to ensure the sample was restricted to those most at risk of developing dementia during the follow-up period. We also further excluded participants with missing APOE e4 carrier status. For T2D, we excluded both prevalent Type 1 diabetes (T1D) and T2D cases at baseline (i.e. UKB enrolment) [2]. The “prevalence algorithm 1” by Eastwood et al. [3] and hospital inpatient records were used to identify prevalent type 1 or type 2 diabetes at baseline.

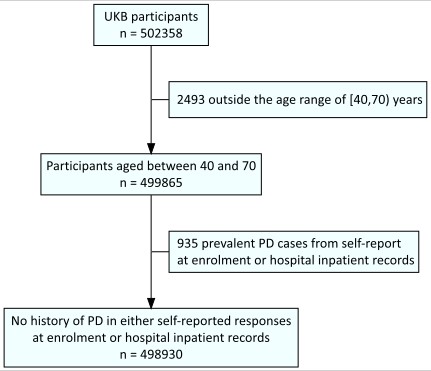

Figure S2. Flowchart of study population for Parkinson's disease

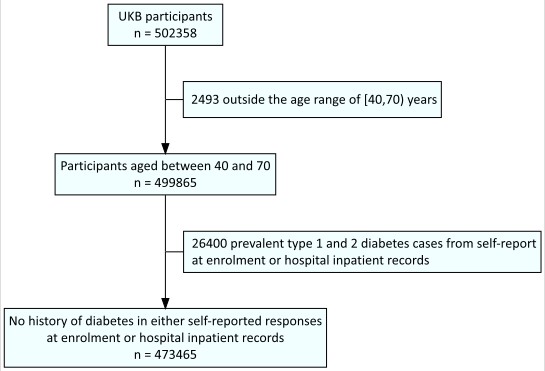

Figure S3. Flowchart of study population for type 2 diabetes

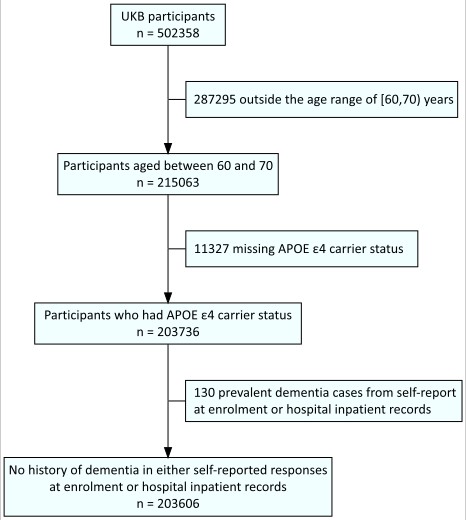

Figure S4. Flowchart of study population for dementia. Note that APEO Ɛ4 is an established risk factor that we included in the risk prediction model for dementia.

#### 3.2 Baseline characteristics for each disease population

*Table S8. Population characteristics for Parkinson’s disease*

|  | **England (N=442577)** | **Scotland (N=35614)** | **Wales**  **(N=20739)** | **Total**  **(N=498930)** |
| --- | --- | --- | --- | --- |
| **Baseline Age (years)** |  |  |  |  |
| Mean (SD) | 56.99 (8.07) | 56.76 (8.03) | 56.63 (7.91) | 56.96 (8.06) |
| Median  (Q1, Q3) | 58.27  (50.52, 63.62) | 57.65  (50.26, 63.50) | 57.73  (50.40, 63.17) | 58.20  (50.50, 63.59) |
| Min - Max | 40.05 - 69.99 | 40.34 - 69.99 | 40.25 - 69.98 | 40.05 - 69.99 |
| Missing | 0 | 0 | 0 | 0 |
| **Sex** |  |  |  |  |
| Female | 240631 (54.4%) | 19827 (55.7%) | 11260 (54.3%) | 271718 (54.5%) |
| Male | 201946 (45.6%) | 15787 (44.3%) | 9479 (45.7%) | 227212 (45.5%) |
| Missing | 0 | 0 | 0 | 0 |
| **Ethnicity** |  |  |  |  |
| White | 414263 (93.6%) | 34910 (98.0%) | 20114 (97.0%) | 469287 (94.1%) |
| Black | 7868 (1.8%) | 56 (0.2%) | 95 (0.5%) | 8019 (1.6%) |
| S. Asian | 7678 (1.7%) | 183 (0.5%) | 115 (0.6%) | 7976 (1.6%) |
| Mixed | 2718 (0.6%) | 89 (0.3%) | 136 (0.7%) | 2943 (0.6%) |
| Other | 7488 (1.7%) | 264 (0.7%) | 197 (0.9%) | 7949 (1.6%) |
| Missing | 2562 (0.6%) | 112 (0.3%) | 82 (0.4%) | 2756 (0.6%) |
| **Townsend Deprivation** |  |  |  |  |
| Q1: least deprived | 87881 (19.9%) | 7986 (22.4%) | 4706 (22.7%) | 100573 (20.2%) |
| Q2 | 89616 (20.2%) | 5969 (16.8%) | 4257 (20.5%) | 99842 (20.0%) |
| Q3 | 87504 (19.8%) | 6198 (17.4%) | 4757 (22.9%) | 98459 (19.7%) |
| Q4 | 88812 (20.1%) | 6903 (19.4%) | 4131 (19.9%) | 99846 (20.0%) |
| Q5: most deprived | 88247 (19.9%) | 8483 (23.8%) | 2858 (13.8%) | 99588 (20.0%) |
| Missing | 517 (0.1%) | 75 (0.2%) | 30 (0.1%) | 622 (0.1%) |
| **Family History of PD** |  |  |  |  |
| No | 425045 (96.0%) | 34314 (96.3%) | 19886 (95.9%) | 479245 (96.1%) |
| Yes | 17532 (4.0%) | 1300 (3.7%) | 853 (4.1%) | 19685 (3.9%) |
| **Smoking status** |  |  |  |  |
| Never | 240546 (54.4%) | 19717 (55.4%) | 11384 (54.9%) | 271647 (54.4%) |
| Previous | 153365 (34.7%) | 11335 (31.8%) | 6928 (33.4%) | 171628 (34.4%) |
| Current | 45997 (10.4%) | 4407 (12.4%) | 2344 (11.3%) | 52748 (10.6%) |
| Missing | 2669 (0.6%) | 155 (0.4%) | 83 (0.4%) | 2907 (0.6%) |

*Table S9. Population characteristics for type 2 diabetes*

|  | **England (N=419839)** | **Scotland**  **(N=34014)** | **Wales**  **(N=19612)** | **Total**  **(N=473465)** |
| --- | --- | --- | --- | --- |
| **Baseline Age (years)** |  |  |  |  |
| Mean (SD) | 56.82 (8.08) | 56.60 (8.04) | 56.45 (7.92) | 56.79 (8.07) |
| Median  (Q1, Q3) | 58.03  (50.28, 63.47) | 57.42  (50.01, 63.35) | 57.43  (50.10, 63.00) | 57.96  (50.26, 63.44) |
| Min - Max | 40.05 - 69.99 | 40.34 - 69.99 | 40.25 - 69.98 | 40.05 - 69.99 |
| Missing | 0 | 0 | 0 | 0 |
| **Ethnicity** |  |  |  |  |
| White | 394916 (94.1%) | 33379 (98.1%) | 19051 (97.1%) | 447346 (94.5%) |
| Other ethnicity | 22598 (5.4%) | 530 (1.6%) | 481 (2.5%) | 23609 (5.0%) |
| Missing | 2325 (0.6%) | 105 (0.3%) | 80 (0.4%) | 2510 (0.5%) |
| **Townsend Deprivation** |  |  |  |  |
| Q1: least deprived | 84475 (20.1%) | 7730 (22.7%) | 4523 (23.1%) | 96728 (20.4%) |
| Q2 | 85799 (20.4%) | 5755 (16.9%) | 4044 (20.6%) | 95598 (20.2%) |
| Q3 | 83331 (19.8%) | 5947 (17.5%) | 4527 (23.1%) | 93805 (19.8%) |
| Q4 | 83816 (20.0%) | 6599 (19.4%) | 3870 (19.7%) | 94285 (19.9%) |
| Q5: most deprived | 81145 (19.3%) | 7859 (23.1%) | 2584 (13.2%) | 91588 (19.3%) |
| Missing | 1273 (0.3%) | 124 (0.4%) | 64 (0.3%) | 1461 (0.3%) |
| **Family History of Diabetes** |  |  |  |  |
| No | 333760 (79.5%) | 28043 (82.4%) | 15419 (78.6%) | 377222 (79.7%) |
| Yes | 86079 (20.5%) | 5971 (17.6%) | 4193 (21.4%) | 96243 (20.3%) |
| **Waist circumference (cm)** |  |  |  |  |
| Mean (SD) | 89.67 (13.09) | 88.04 (12.87) | 90.94 (13.20) | 89.61 (13.09) |
| Median  (Q1, Q3) | 89.00  (80.00, 98.00) | 88.00  (78.00, 96.00) | 91.00  (81.00, 100.00) | 89.00  (80.00, 98.00) |
| Min - Max | 20.00 - 182.00 | 20.00 - 171.00 | 52.00 - 160.00 | 20.00 - 182.00 |
| Missing | 1775 (0.4%) | 103 (0.3%) | 79 (0.4%) | 1957(0.4%) |
| **BMI** |  |  |  |  |
| Underweight/Normal | 143748 (34.2%) | 11711 (34.6%) | 5767 (29.4%) | 161226 (34.3%) |
| Overweight | 178930 (42.6%) | 14510 (42.7%) | 8485 (43.3%) | 201925 (42.6%) |
| Obese | 94661 (22.5%) | 7654 (22.5%) | 5242 (26.7%) | 107557 (22.7%) |
| Missing | 2500 (0.6%) | 139 (0.4%) | 118 (0.6%) | 2757 (0.6%) |
| **Physical inactivity** |  |  |  |  |
| No | 178826 (42.6%) | 14027 (41.2%) | 7862 (40.1%) | 200715 (42.4%) |
| Yes | 144163 (34.3%) | 12426 (36.5%) | 7131 (36.4%) | 163720 (34.6%) |
| Missing | 96850 (23.1%) | 7561 (22.2%) | 4619 (23.6%) | 109030 (23.0%) |
| **Hypertension** |  |  |  |  |
| No | 192546 (45.9%) | 14689 (43.2%) | 8237 (42.0%) | 215472 (45.5%) |
| Yes | 227293 (54.1%) | 19325 (56.8%) | 11375 (58.0%) | 257993 (54.5%) |

*Table S10. Population characteristics for dementia*

|  | **England (N=181850)** | **Scotland (N=13773)** | **Wales**  **(N=7983)** | **Total (N=203606)** |
| --- | --- | --- | --- | --- |
| **Baseline Age (years)** |  |  |  |  |
| Mean (SD) | 64.56 (2.81) | 64.72 (2.90) | 64.40 (2.73) | 64.56 (2.81) |
| Median  (Q1, Q3) | 64.36  (62.13, 66.84) | 64.62  (62.08, 67.21) | 64.19  (62.02, 66.68) | 64.37  (62.12, 66.86) |
| Min - Max | 60.00 - 69.99 | 60.00 - 69.99 | 60.00 - 69.98 | 60.00 - 69.99 |
| Missing | 0 | 0 | 0 | 0 |
| **Ethnicity** |  |  |  |  |
| White | 175391 (96.4%) | 13615 (98.9%) | 7844 (98.3%) | 196850 (96.7%) |
| Other ethnicity | 5568 (3.1%) | 121 (0.9%) | 114 (1.4%) | 5803 (2.9%) |
| Missing | 891 (0.5%) | 37 (0.3%) | 25 (0.3%) | 953 (0.5%) |
| **Townsend Deprivation** |  |  |  |  |
| Q1: least deprived | 36821 (20.2%) | 3081 (22.4%) | 1811 (22.7%) | 41713 (20.5%) |
| Q2 | 37453 (20.6%) | 2264 (16.4%) | 1637 (20.5%) | 41354 (20.3%) |
| Q3 | 34577 (19.0%) | 2306 (16.7%) | 1746 (21.9%) | 38629 (19.0%) |
| Q4 | 31705 (17.4%) | 2372 (17.2%) | 1317 (16.5%) | 35394 (17.4%) |
| Q5: most deprived | 27393 (15.1%) | 2789 (20.2%) | 815 (10.2%) | 30997 (15.2%) |
| Missing | 13901 (7.6%) | 961 (7.0%) | 657 (8.2%) | 15519 (7.6%) |
| **Family History of Dementia** |  |  |  |  |
| No | 154574 (85.0%) | 11791 (85.6%) | 6942 (87.0%) | 173307 (85.1%) |
| Yes | 27276 (15.0%) | 1982 (14.4%) | 1041 (13.0%) | 30299 (14.9%) |
| **Smoking status** |  |  |  |  |
| Never | 90237 (49.6%) | 6830 (49.6%) | 3988 (50.0%) | 101055 (49.6%) |
| Previous | 75973 (41.8%) | 5355 (38.9%) | 3237 (40.5%) | 84565 (41.5%) |
| Current | 14525 (8.0%) | 1515 (11.0%) | 724 (9.1%) | 16764 (8.2%) |
| Missing | 1115 (0.6%) | 73 (0.5%) | 34 (0.4%) | 1222 (0.6%) |
| **APOE_e4_carrier** |  |  |  |  |
| Not a carrier | 133893 (73.6%) | 10047 (72.9%) | 5977 (74.9%) | 149917 (73.6%) |
| e4 carrier | 47957 (26.4%) | 3726 (27.1%) | 2006 (25.1%) | 53689 (26.4%) |
| Missing | 0 | 0 | 0 | 0 |
| **Education** |  |  |  |  |
| Below GCSE | 51384 (28.3%) | 4324 (31.4%) | 2305 (28.9%) | 58013 (28.5%) |
| Equivalent or above  GCSE | 127817 (70.3%) | 9254 (67.2%) | 5565 (69.7%) | 142636 (70.1%) |
| Missing | 2649 (1.5%) | 195 (1.4%) | 113 (1.4%) | 2957 (1.5%) |
| **physical inactivity** |  |  |  |  |
| No | 78716 (43.3%) | 5702 (41.4%) | 3145 (39.4%) | 87563 (43.0%) |
| Yes | 57071 (31.4%) | 4522 (32.8%) | 2650 (33.2%) | 64243 (31.6%) |
| Missing | 46063 (25.3%) | 3549 (25.8%) | 2188 (27.4%) | 51800 (25.4%) |
| **Depression** |  |  |  |  |
| No | 122619 (67.4%) | 9237 (67.1%) | 5334 (66.8%) | 137190 (67.4%) |
| Yes | 58730 (32.3%) | 4511 (32.8%) | 2631 (33.0%) | 65872 (32.4%) |
| Missing | 501 (0.3%) | 25 (0.2%) | 18 (0.2%) | 544 (0.3%) |
| **Diabetes** |  |  |  |  |
| No | 168614 (92.7%) | 12858 (93.4%) | 7352 (92.1%) | 188824 (92.7%) |
| Yes | 12960 (7.1%) | 905 (6.6%) | 625 (7.8%) | 14490 (7.1%) |
| Missing | 276 (0.2%) | 10 (0.1%) | 6 (0.1%) | 292 (0.1%) |
| **Hearing impairment** |  |  |  |  |
| No | 123903 (68.1%) | 9772 (71.0%) | 5398 (67.6%) | 139073 (68.3%) |
| Yes | 56164 (30.9%) | 3812 (27.7%) | 2497 (31.3%) | 62473 (30.7%) |
| Missing | 1783 (1.0%) | 189 (1.4%) | 88 (1.1%) | 2060 (1.0%) |
| **Hypertension** |  |  |  |  |
| No | 53864 (29.6%) | 3602 (26.2%) | 2052 (25.7%) | 59518 (29.2%) |
| Yes | 127986 (70.4%) | 10171 (73.8%) | 5931 (74.3%) | 144088 (70.8%) |
| **Social isolation** |  |  |  |  |
| No | 165581 (91.1%) | 12505 (90.8%) | 7306 (91.5%) | 185392 (91.1%) |
| Yes | 16049 (8.8%) | 1258 (9.1%) | 672 (8.4%) | 17979 (8.8%) |
| Missing | 220 (0.1%) | 10 (0.1%) | 5 (0.1%) | 235 (0.1%) |
| **BMI** |  |  |  |  |
| Underweight/Normal | 54477 (30.0%) | 4064 (29.5%) | 2000 (25.1%) | 60541 (29.7%) |
| Overweight | 81831 (45.0%) | 6202 (45.0%) | 3624 (45.4%) | 91657 (45.0%) |
| Obese | 44803 (24.6%) | 3469 (25.2%) | 2318 (29.0%) | 50590 (24.8%) |
| Missing | 739 (0.4%) | 38 (0.3%) | 41 (0.5%) | 818 (0.4%) |

#### 3.3 Comparisons via Cox models

For Cox models, prevalent cases identified by (i) self-report (diagnoses and medications) UKB data at enrolment date and (ii) hospital inpatient data prior to or at enrolment were excluded in analysis. Incident cases were ascertained longitudinally using record-level hospital inpatient data and death registry. The sources for disease ascertainment are the same as in Clifton *et. al.* (2024). A follow-up time of each participant was calculated as the number of years from the date of baseline assessment until the earliest date among the following: date of the first diagnosis for the specific disease, death date by other causes, loss to follow-up date, or the earliest UKB administrative censoring date among the three countries (31 May 2022).

Established risk factors for each disease were identified from the literature [4]–[8], with full detail published in the supplementary materials by Clifton et al. [9]. We kept the derivation and categorisation of risk factors consistent across the diseases wherever possible. For example, we used BMI “underweight/normal, overweight, and obese” consistently. Our “country” variable is derived using the assessment centre (UK Biobank Data-Field 54) at baseline.

*Table S11. Cox models for Parkinson’s disease (N=498,930). 95% CI: 95% confidence interval. Model 1: age + sex + self-reported ethnicity + Townsend deprivation score quintile + Family history of PD + smoking; Model 2: Model 1 + country*

|  | **Model 1** | **Model 2** |
| --- | --- | --- |
|  | **HR (95% CI, p-value)** | **HR (95% CI, p-value)** |
| **Age at enrolment (Year)** | 1.14 (1.13-1.15, p<0.001) | 1.14 (1.13-1.15, p<0.001) |
| **Sex (Reference group: Female)** |  |  |
| Male | 1.90 (1.78-2.03, p<0.001) | 1.90 (1.78-2.03, p<0.001) |
| **Ethnicity (Reference group: White)** |  |  |
| Other ethnicity | 1.07 (0.90-1.26, p=0.452) | 1.04 (0.88-1.22, p=0.673) |
| **Townsend Deprivation Quintile (Reference group: Q1)** |  |  |
| Q2 | 0.96 (0.87-1.07, p=0.479) | 0.96 (0.87-1.06, p=0.376) |
| Q3 | 1.02 (0.92-1.12, p=0.749) | 1.01 (0.91-1.12, p=0.832) |
| Q4 | 1.07 (0.97-1.19, p=0.179) | 1.07 (0.97-1.18, p=0.201) |
| Q5 (most deprived) | 1.26 (1.14-1.40, p<0.001) | 1.27 (1.14-1.40, p<0.001) |
| **Family history of PD (Reference group: No)** |  |  |
| Yes | 1.73 (1.53-1.95, p<0.001) | 1.73 (1.53-1.95, p<0.001) |
| **Smoking Status (Reference group: Never)** |  |  |
| Previous | 0.93 (0.87-1.00, p=0.046) | 0.93 (0.87-0.99, p=0.034) |
| Current | 0.78 (0.68-0.88, p<0.001) | 0.78 (0.69-0.89, p<0.001) |
| **Country (Reference group: England):** |  |  |
| Scotland | NA | 0.62 (0.54-0.72, p<0.001) |
| Wales | NA | 0.77 (0.64-0.92, p=0.004) |

*Table S12. Cox models for type 2 diabetes (T2D) (N=473,465). 95% CI: 95% confidence interval.* ***Model 1****: age + self-reported ethnicity + Townsend deprivation score quintile + family history of T2D + waist circumference (cm) + BMI + physical inactivity + hypertension;* ***Model 2****: Model 1 + country*

|  | **Model 1** | **Model 2** |
| --- | --- | --- |
|  | **HR (95% CI, p-value)** | **HR (95% CI, p-value)** |
| **Age at enrolment (Year)** | 1.05 (1.05-1.05, p<0.001) | 1.05 (1.05-1.05, p<0.001) |
| **Ethnicity (Reference group: White)** |  |  |
| Other ethnicity | 2.37 (2.24-2.51, p<0.001) | 2.29 (2.16-2.43, p<0.001) |
| **Townsend Deprivation Quintile (Reference group: Q1)** |  |  |
| Q2 | 1.09 (1.03-1.16, p=0.002) | 1.08 (1.02-1.14, p=0.005) |
| Q3 | 1.14 (1.08-1.21, p<0.001) | 1.14 (1.08-1.20, p<0.001) |
| Q4 | 1.25 (1.18-1.32, p<0.001) | 1.25 (1.18-1.32, p<0.001) |
| Q5 (most deprived) | 1.64 (1.56-1.73, p<0.001) | 1.66 (1.57-1.74, p<0.001) |
| **Family history of diabetes (Reference group: No)** |  |  |
| Yes | 1.84 (1.78-1.91, p<0.001) | 1.84 (1.77-1.90, p<0.001) |
| **Waist (cm)** | 1.05 (1.04-1.05, p<0.001) | 1.04 (1.04-1.05, p<0.001) |
| **BMI category (Reference group: Underweight/Normal)** |  |  |
| Overweight | 1.58 (1.49-1.68, p<0.001) | 1.59 (1.50-1.70, p<0.001) |
| Obese | 2.32 (2.16-2.49, p<0.001) | 2.36 (2.20-2.53, p<0.001) |
| **Physical inactivity (Reference group: No)** |  |  |
| Yes | 1.15 (1.11-1.19, p<0.001) | 1.15 (1.12-1.19, p<0.001) |
| **Hypertension (Reference group: No)** |  |  |
| Yes | 1.70 (1.63-1.77, p<0.001) | 1.71 (1.64-1.78, p<0.001) |
| **Country (Reference group: England):** |  |  |
| Scotland | NA | 0.49 (0.45-0.54, p<0.001) |
| Wales | NA | 0.96 (0.89-1.04, p=0.328) |

*Table S13. Cox models for dementia (N=203,606). 95% CI: 95% confidence interval. Model 1:* *age + sex + self-reported ethnicity + Townsend deprivation score quintile + Family history of dementia + smoking + APOE_e4_carrier + education + physical inactivity + depression + diabetes + hearing impairment + hypertension + social isolation + BMI;* *Model 2:* *Model 1 + country*

|  | **Model 1** | **Model 2** |
| --- | --- | --- |
|  | **HR (95% CI, p-value)** | **HR (95% CI, p-value)** |
| **Age at enrolment (Year)** | 1.24 (1.22-1.25, p<0.001) | 1.24 (1.22-1.25, p<0.001) |
| **Sex (Reference group: Female)** |  |  |
| Male | 1.26 (1.18-1.34, p<0.001) | 1.26 (1.18-1.34, p<0.001) |
| **Ethnicity (Reference group: White)** |  |  |
| Other ethnicity | 1.33 (1.08-1.63, p=0.007) | 1.32 (1.07-1.62, p=0.009) |
| **Townsend Deprivation Quintile (Reference group: Q1)** |  |  |
| Q2 | 0.98 (0.89-1.08, p=0.686) | 0.98 (0.88-1.08, p=0.643) |
| Q3 | 1.02 (0.92-1.13, p=0.677) | 1.02 (0.92-1.13, p=0.705) |
| Q4 | 1.16 (1.05-1.29, p=0.003) | 1.16 (1.05-1.29, p=0.003) |
| Q5 (most deprived) | 1.29 (1.16-1.43, p<0.001) | 1.29 (1.16-1.43, p<0.001) |
| **Family history of dementia (Reference group: No)** |  |  |
| Yes | 1.56 (1.45-1.68, p<0.001) | 1.56 (1.45-1.68, p<0.001) |
| **Smoking status (Reference group: Never)** |  |  |
| Previous | 1.13 (1.05-1.21, p<0.001) | 1.13 (1.05-1.21, p=0.001) |
| Current | 1.36 (1.21-1.52, p<0.001) | 1.36 (1.21-1.53, p<0.001) |
| **APOE_e4_carrier (Reference group: No)** |  |  |
| Yes | 3.55 (3.33-3.79, p<0.001) | 3.55 (3.33-3.79, p<0.001) |
| **Education (Reference group: No)** |  |  |
| Yes | 0.74 (0.69-0.79, p<0.001) | 0.74 (0.69-0.79, p<0.001) |
| **Physical inactivity (Reference group: No)** |  |  |
| Yes | 1.07 (1.00-1.14, p=0.045) | 1.07 (1.00-1.14, p=0.043) |
| **Depression (Reference group: No)** |  |  |
| Yes | 1.25 (1.17-1.34, p<0.001) | 1.25 (1.17-1.34, p<0.001) |
| **Diabetes (Reference group: No)** |  |  |
| Yes | 2.26 (1.22-4.21, p=0.010) | 2.24 (1.20-4.17, p=0.011) |
| **Hearing loss (Reference group: No)** |  |  |
| Yes | 1.07 (1.00-1.14, p=0.053) | 1.07 (1.00-1.14, p=0.058) |
| **Hypertension (Reference group: No)** |  |  |
| Yes | 1.20 (1.11-1.29, p<0.001) | 1.20 (1.11-1.29, p<0.001) |
| **Social isolation (Reference group: No)** |  |  |
| Yes | 1.40 (1.26-1.55, p<0.001) | 1.40 (1.26-1.55, p<0.001) |
| **BMI category (Reference group: Normal)** |  |  |
| Overweight | 0.82 (0.76-0.88, p<0.001) | 0.82 (0.76-0.88, p<0.001) |
| Obese | 0.86 (0.79-0.94, p=0.001) | 0.86 (0.79-0.94, p=0.001) |
| **Country (Reference group: England):** |  |  |
| Scotland | NA | 0.88 (0.78-0.99, p=0.034) |
| Wales | NA | 0.98 (0.83-1.16, p=0.827) |

### 4. Further Discussion

We used the variable “UKB Assessment Centre” to derive the country of residence of each individual at enrollment. We did not take into account the small number of participants who may have moved across the three countries after enrollment and hence have a hospital diagnosis in a different country. We anticipate this number to be small, and thus have negligible influence on the conclusions of this study.
